# An Estimation of Reproduction Number of SARS-CoV-2 by Age Class for Age Classes in Japan

**DOI:** 10.1101/2021.01.14.21249854

**Authors:** Junko Kurita, Takahide Hata, Tamie Sugawara, Yasushi Ohkusa, Atsuko Hata

## Abstract

**Background:** Infectiousness of COVID-19 by age class inferred from the infection source and by infected people might be different. However, studies of such infectiousness have not been reported.

**Object:** The object of this study was estimation of reproduction numbers by age class of the source of infection and the infected persons. To do so, after examining a new procedure for reproduction number estimation, we checked infected places by age class of the source of infection and the infected persons.

**Method:** We ignore patients who infected no one because their reliability might be lower than that of patients who infected more than one person. We estimated the reproduction number from the histogram of the number of the infected people by the same patient, assuming that the histogram follows an exponential distribution.

**Discussion and Conclusion:** The obtained results demonstrated that the effective reproduction numbers for infection from children were very low. They were higher among adult and elderly people than among the same age class. Moreover, although the highest and second-highest infected places were ‘other’ and ‘at home’ with some exceptions, the data for infection at hospitals were remarkable among adults and elderly people. Among elderly people, infection at facilities for elderly was also high. Infections at nursing schools, schools, restaurants, and entertainment venues at night were negligible.

## Introduction

Since the emergence COVID-19 in December, 2019 in Wuhan, China, several reproduction numbers have been estimated. Some of the earliest studies in Wuhan [1–3] estimated R_0_ for COVID-19 as 2.24–3.58. Even in Japan, early research [4] estimated *R*_*0*_ as 2.049 (95%CI [2.403, 2.557]). However, these were reproduction numbers for the whole population. Reproduction numbers by age class probably were lesser known. One exceptional report described the effective reproduction numbers for children as 1.75, 1.84 for adult, and 2.19 for elderly people [5].

However, infectiousness by age class by the source of infection and the infected persons might be different. For example, children might have high infectiousness to children, but low infectiousness to elderly people. Conversely adults might have high infectiousness to elderly people. When considering the effects of school closure as a countermeasure against COVID-19, the most important parameter should be infectiousness among children. Infectiousness of children to adults including teachers and caregivers is less interesting. Therefore, this study was conducted to estimate the reproduction number by age class to the same and other age classes.

An exemplary study conducted in the USA reported that child patients with symptoms shed the virus as much or more than adult patients did [6]. That finding suggests that infectiousness from symptomatic children to adults is comparable to that from one adult to another adult. A report of a study conducted in Japan showed that non-symptomatic nursery school children infected other children or teachers [7]. This finding suggests that infectiousness from symptomatic children to children was almost comparable to infectiousness to adults.

This study was undertaken to estimate reproduction numbers by the age class of the source of infection and of the infected persons. To make such an estimation, we ignore patients who did not infect others at all because their reliability might be lower than that of patients who infected more than one person. Subsequently, we examined new procedures for estimating the reproduction numbers. Moreover, we checked the infected places by age class of the infection source and the infected persons.

## Method

Let *x*_*i,j,k*_ represent the number of cases in which *k* patient in *i* age class were infected from the same patient who was of *j* age class. Particularly, we presumed three age classes: *i* or *j*=1 represents an age class of younger than 20 years old; 2 indicates the age class of patients who were 20 years old or older and younger than 60 years old; and 3 indicates the age class of people 60 years old and older. Additionally, *y*_*i*,_ represents the total number of patients including those for whom the source of infection was unidentified. Therefore, *R*_*i,j*_, which represents the effective reproduction number from age class *j* to age class *i*, was defined as *Σ*_*k*=1_ *k x*_*i,j,k*_ /*y*_*j*_.

However, the sources of infection of almost all patients were unidentified. Actually, the source of infection was unknown in 85% of infected people of all age classes. Therefore, *Σ*_*k*=*1*_ *x*_*i,j,k*_ is expected to be much less than *y*_*j*_. In fact, these patients were infected by someone, even if their source of infection was not identified. Moreover, if a patient was not confirmed as infecting anyone (nobody confirmed being infected by a patient), it cannot be said that this patient did not actually infect anyone. Therefore, unidentified sources of infection or persons not confirmed as having infected anyone were less reliable than cases in which the source of infection had been confirmed. Therefore, we limited the information to be used to about *x*_*i,j,k*_ for *k*>*0*.

Because we do not know the probability that a patient infected one person, the probability that a person infecting two or more people was assumed to follow an exponential distribution as 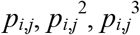, and so on. Then 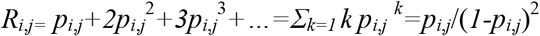.

We observed an estimator of *p*_*i,j*_ as *x*_*i,j*,1_/*N*_*i,j*_, where *N*_*i,j*_ was an unknown total number of age class *i* patients who were infected by age class *j* patients. Similarly, 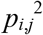 was estimated as *x*_*i,j*,2_/*N*_*i,j*_ and 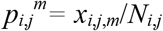 in general. By log transformation, we have *m log p*_*i,j*_ = *logx*_*i,j,m*_*log N*_*i,j*_ (*m*= 1,2,…). Therefore, we obtain an estimator of *p* as an estimated coefficient of regression of *log x*_*i,j,m*_ on *m* by ordinary least squares method. In addition, 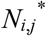 was obtained from an exponential transform for the estimated constant term.

The confidence interval (CI) of 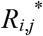 was obtained using a bootstrapping procedure for the distribution of {*x*_*i,j,m*_ (*m*=1,2,…)}. Based on the *k-th* bootstrapped distribution, {*x*_*i,j,m*_ (*m*=1,2,…)}^*k*^, we can obtain 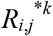. We repeated this procedure one million times, thereby obtaining one million bootstrapped 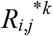. We sorted these variables. The duration from 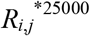 to 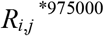 is expected to be 95%CI of 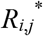.

In addition to that, we classified the used data to the infection place by age class of the source of infection and the infected. The infected places which were classified were homes, hospitals, facilities for elderly people, schools except for universities, nursery schools, universities, restaurants other than entertainment at night or karaoke, entertainment at night, karaoke, workplaces, sports gym, theaters and live venues, others, and unknown.

All information used for this study was obtained from reports of the Ministry of Health, Labour and Welfare [8] and local governments. The study period extended from January 15, when the initial case was detected in Japan, to the end of July.

## Ethical considerations

All information used for this study has been published elsewhere [8]. There is therefore no ethical issue related to this study.

## Results

Through the end of July, 36,431 patients had been confirmed in Japan. Of them, excluding asymptomatic cases, cases of people presumed to have been infected in foreign countries, and cases whose onset date were not available, we are left with 30,780 cases. Of those cases, excluding cases for which the source of infection was unknown, and for which the age of patients themselves and source of infection were unavailable, we are left with 5383 cases. Of those, 4886 cases were identified as the infection source. These 4886 cases were analyzed for this study.

Estimation results of *R*_*i,j*_ are presented in Table 1. High reproduction numbers were found among adults and elderly people: higher than 4. Moreover, among elderly people and among adults, *R*_*i,j*_ were, respectively, approximately 3.7 and 2.5. Conversely, to children from adults or elderly people, *R*_*i,j*_ were very small: approximately 0.3 or 0.1, respectively. From children, *R*_*i,j*_ were less than one. The highest *R*_*i,j*_ was to adults, but it was less than 0.7.

**Table 1:** Estimate results of *R*_*i,j*_.

| Source of Infection: Children |  |  |  |
| --- | --- | --- | --- |
| Infected | Estimated | 95%CI |  |
|  |  | Lower bound | Upper bound |
| Children | 0.6037 | 0.4793 | 0.7465 |
| Adult | 0.6221 | 0.4931 | 0.7604 |
| Elderly | 0.1244 | 0.0783 | 0.1797 |
| Source of Infection: Adult |  |  |  |
| Children | 0.2709 | 0.2608 | 0.2812 |
| Adult | 2.4944 | 2.4436 | 2.5454 |
| Elderly | 4.5258 | 4.4449 | 4.6076 |
| Source of Infection: Elderly |  |  |  |
| Children | 0.1251 | 0.1134 | 0.1370 |
| Adult | 4.9616 | 4.8041 | 5.1220 |
| Elderly | 3.7169 | 3.5898 | 3.8464 |

Note: Children were defined as patients with age younger than 20 years old. Adult was defined as patients with age older than or was 20 years old and younger than 60 years old. Elderly was defined as patients 60 years old and older.

Table 2 presents infection places in the data considered for this study. It shows that “others” were consistently the highest proportion among any combination of age class of source of infection and the infected persons. The second highest were at “home”, except for infections from children to children. The second highest proportion of infection place from children to children was “school”, which accounted for about 30%. The third highest proportion among adult and elderly people was at a “hospital”. However, among elderly people, the third highest was at “facilities for elderly people”. Among adults, the third highest was at the “workplace”. From children to adults, it was at a “school or university”. No infection was recorded at a “nursery school” among children; less than 1% of all infections were from children to adults. Infection at “restaurants” was less than 2% among adults. “Entertainment at night venues” accounted for less than 0.4% of infections among adults and among elderly people.

**Table 2:**
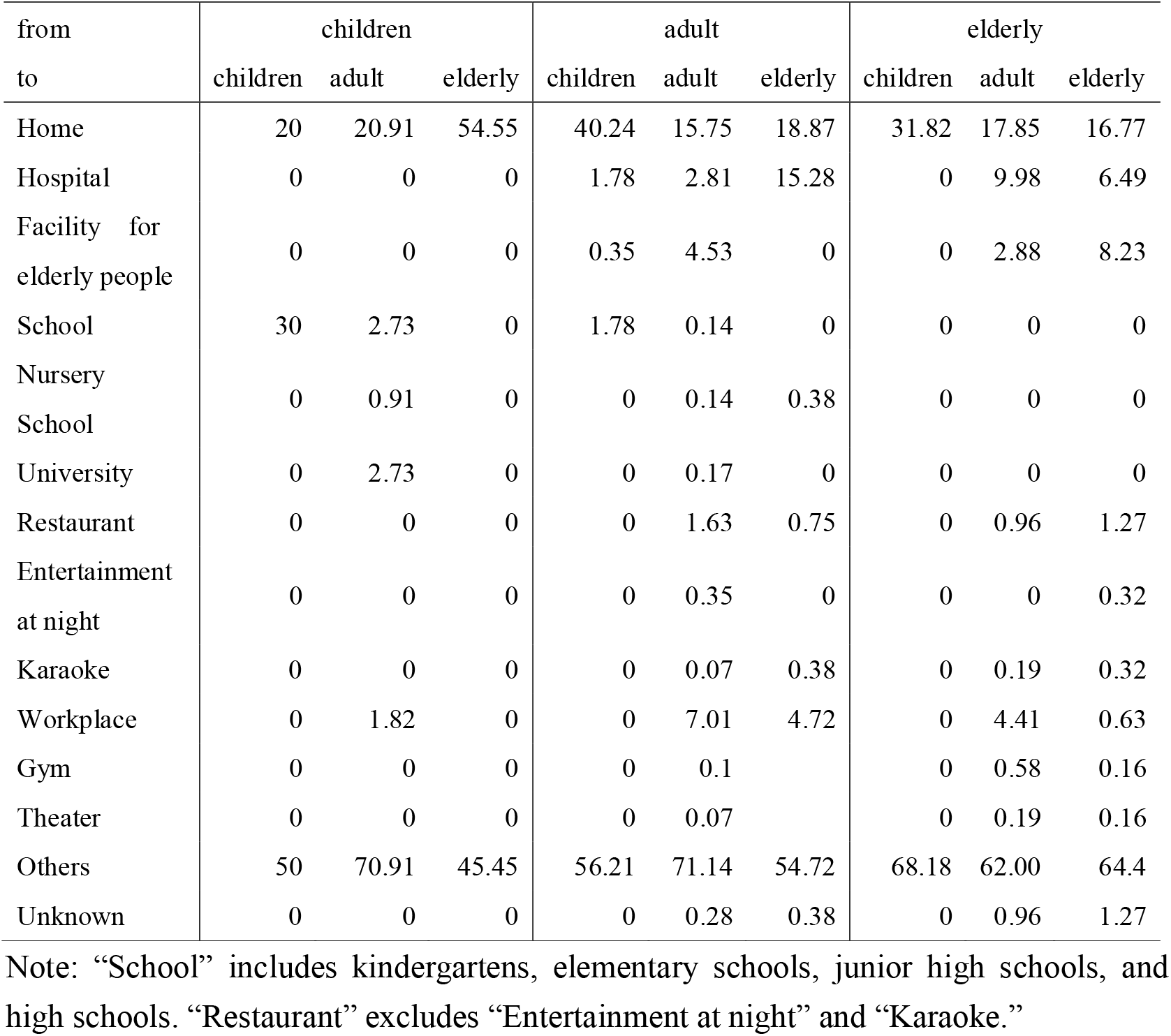
Distribution of cases over infected place by age class of source of infection and infected persons among patients whose source of infection were identified (%)

## Discussion

We developed a procedure to estimate case distributions among the numbers of the infected cases. It ignores information about infected cases or unlinked cases for which the source of infection was unknown, although they were a majority because their information was less credible. However, information about patients who were reported as having infected someone was more reliable than others because, at least, they were investigated by public health authorities.

Earlier studies have included patients who were not reported as having infected someone [7]. They estimated a very small reproduction number, 0.6, as of the end of February in Japan. Although they did not call it *R*_*0*_, they referred to it as the average of the secondary infection. That number indicates that the outbreak of COVID-19 was self-limited. Therefore, any intensive infection control such as school closure or restriction against going out is expected to be unnecessary. The authors of that report apparently misunderstood the meaning of patients who were not reported as having infected someone. They might have been under-investigated at that time. Therefore, the people infected by them might have been found and reported. Alternatively, investigation of them simply cannot find out who had been infected by them.

When we applied our procedure used in the present study to the data of an earlier study, we obtained a figure of 4.4273; its 95%CI was [3.6000, 5.3364]. That value is more than six times larger than the original estimate. It was almost comparable with our obtained results for infections from adult/elderly to elderly/adult. They apparently underestimated *R*_*0*._ Therefore, they misled infection-control policy and insisted on contact tracing.

Results demonstrated that children are unlikely to infect anyone, even with *R*_*i,j*_>1. Apparently, they cannot spread the virus effectively. In this sense, school or class closure might be less meaningful because an outbreak in a class or school might be a lower possibility. In contrast, *R*_*i,j*_ from adult/elderly to elderly/adult were very high. They were presumed to contact at home or at facilities for elderly people. Precautions at these places are expected to be the most important for infection control.

Our obtained results, especially among adults and elderly people, are apparently higher than estimation of reproduction number of COVID-19 for the entire population to date. However, infectiousness to an age class by an age class has not been explored. We cannot judge that these were overestimations through comparison with earlier studies.

One possible reason why overestimation occurred might be the small sample used for estimation. As described above, the total number of data was less than 5000 patients. Therefore, a histogram of the number of secondary infections should be scarce sometimes, especially in cases for which the infection source was children. If using more data, then the estimation might improve. However, it is noteworthy that larger amounts of data for adult and elderly people than for children lead to estimates that were much larger.

Another potential reason might be estimation procedures. Especially, there were outliers for which the number of secondary infections was larger than 20. They certainly affected estimation strongly. They might be so-called “super spreaders,” although their credibility might be lower than that of non-super-spreaders. However, some infected people were reported. As shown in Figures 2 and 3, they were observed only among adult and elderly people. Therefore, they were presumed to be infected at a facility such as a hospital or a facility for elderly people. To allay their effect, we can use robust estimation instead of ordinary least squares in a study. It remains as a challenge for future research.

**Figure 1:**
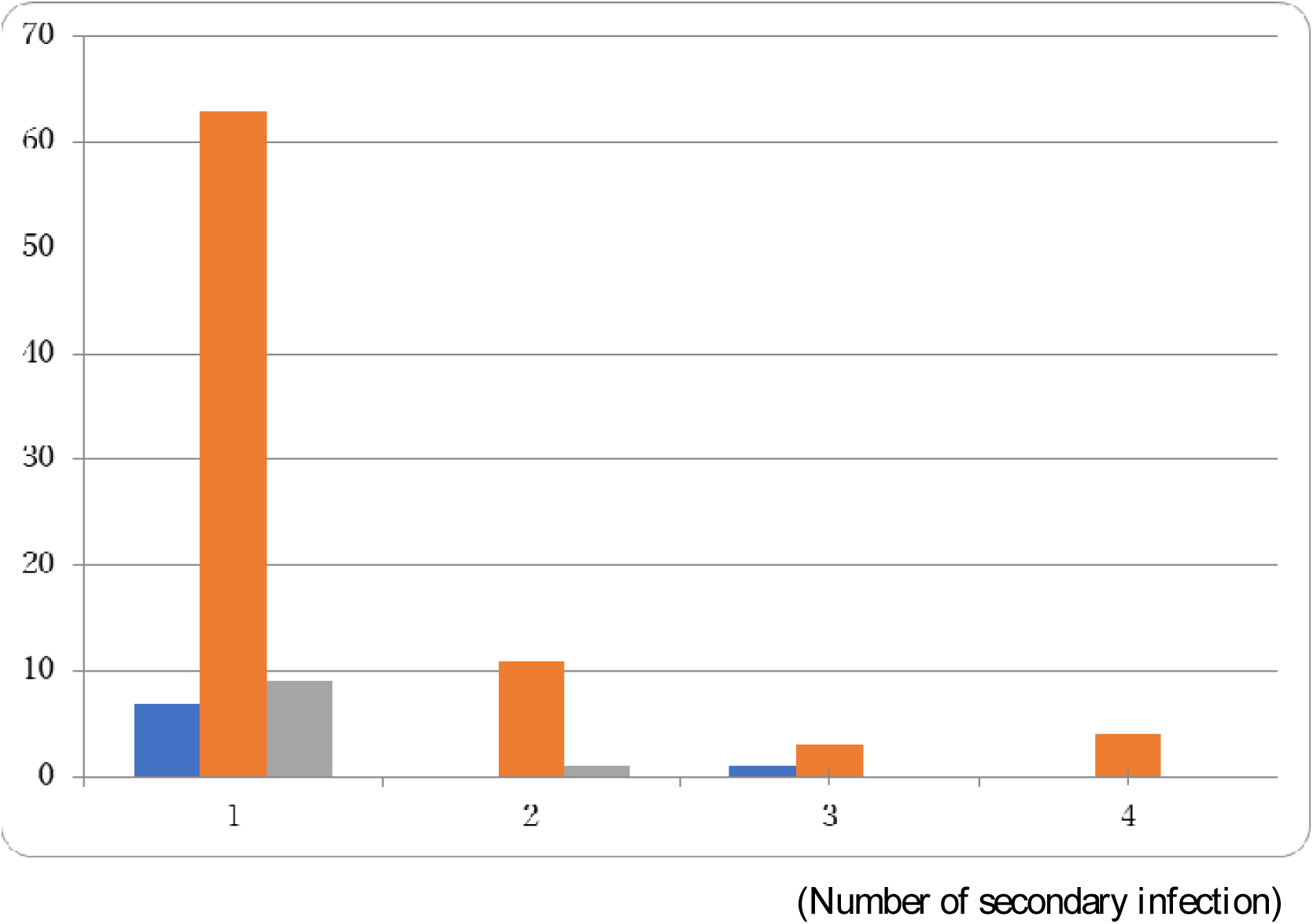
Histogram of the number of cases from children by number of secondary infection. Note: Blue bars represent the number of cases attributable to children, red bars represent cases attributable to adult, and green bars represent cases attributable to elderly people.

**Figure 2:**
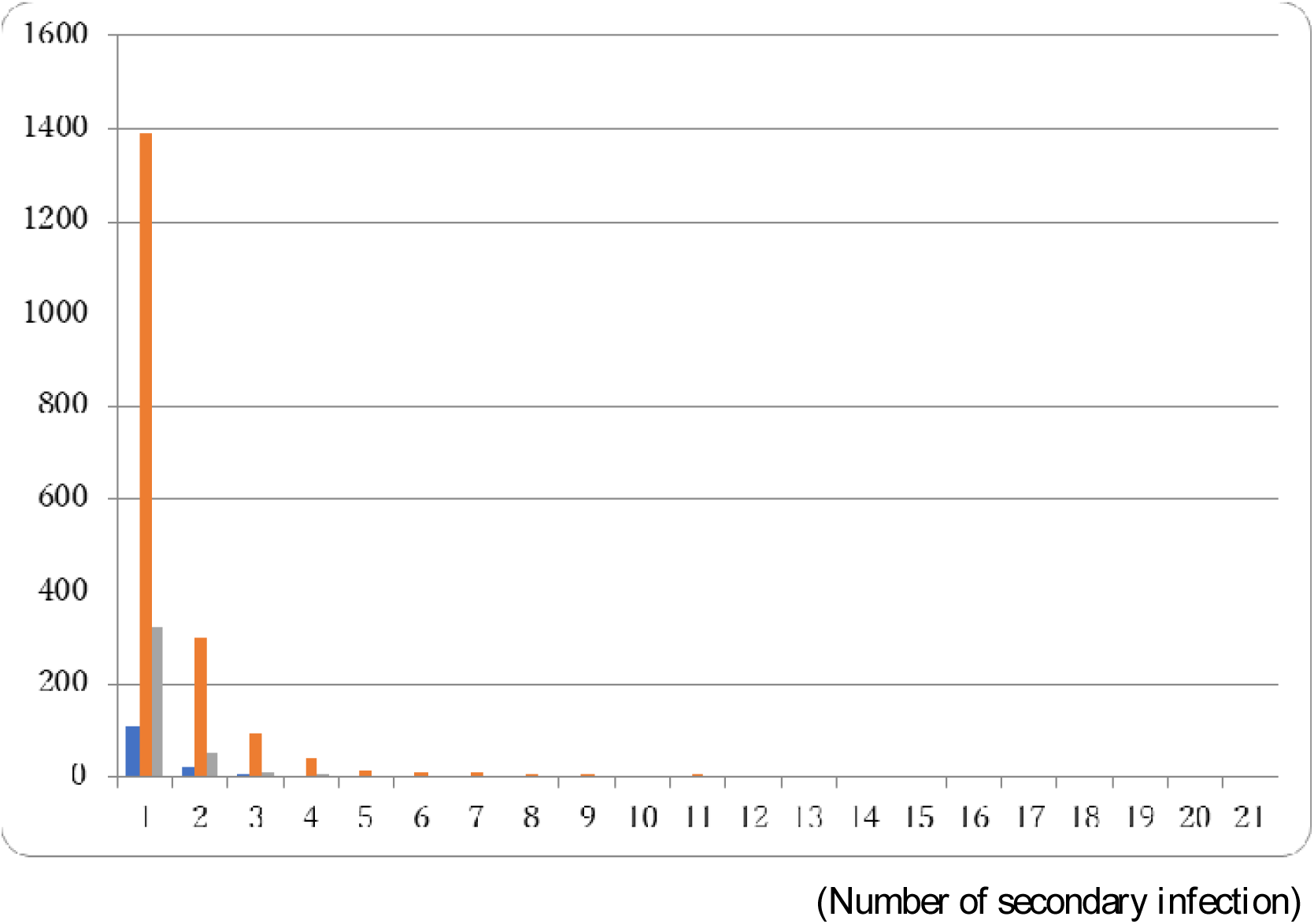
Histogram of the number of cases attributable to adults by the number of secondary infections. Note: Blue bars represent the number of cases attributable to children, red bars represent cases attributable to adult, and green bars represent cases attributable to elderly people.

**Figure 3:**
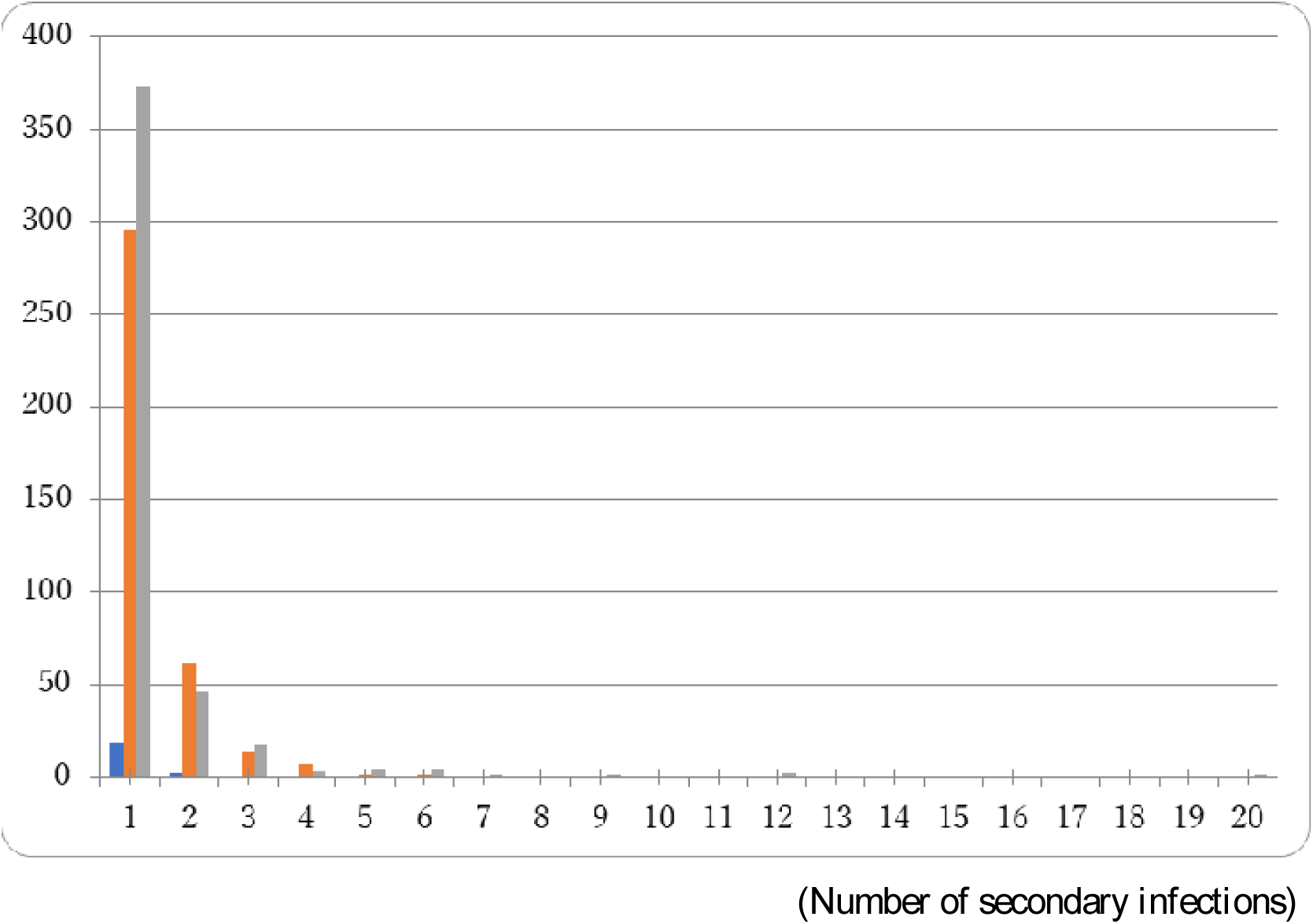
Histogram of the number of cases attributable to infection by elderly people by the number of secondary infections. Note: Blue bars represent the number of cases attributable to children, red bars represent cases attributable to adult, and green bars represent cases attributable to elderly people.

From the distribution of infected places by age class of source of infection and the infected, we can learn many points about COVID-19 characteristics and countermeasures against it. First, because others were consistently the highest proportion among combinations of age classes of infection sources and infected people, there were many types of infected places: it was difficult to isolate an infection place. Taking the source of infection as occupying about 60% consistently into consideration, countermeasures for any particular place including restaurants or entertainment nightspots might be meaningless. These locations were sometimes regulated for application of countermeasures in Japan, but their proportions were just less than 2% and 0.4%, respectively. Therefore, the effectiveness of those regulations might be weak.

Secondly, because the second highest proportion were those infected at home, countermeasures at home might be important, except for infection from children to children. Nevertheless, we cannot shut down homes. The effectiveness of precautions at homes might be limited.

Thirdly, the third highest proportion of infection among adult and elderly people was at hospital and was at facilities for elderly among elderly people. Clearly, they were vulnerable to COVID-19. Precautions at hospitals or facilities for elderly people have been quite important.

Fourthly, among children, the second highest proportion of infection was school. However, it was very rare. Infectiousness among children was low. Moreover, there was no infection at nursery schools among children and just a less than 1% of infection from children to adults. Schools and nursery schools might not be a locus of infection as they are for influenza or other pediatric infectious diseases.

The present study has some limitations. First, we used data that were limited to those for which the infection source was identified, with the age class of the infected persons reported, and infected people all with some symptoms. However, those symptomatic patients examined for the present study were very rare among all symptomatic patients. Sources of infection for most of patients were unknown and might not be reported in public even after being identified by the public health center. Therefore, we assumed implicitly that they were representable in all symptomatic patients. The data validity should be confirmed.

Second, similar to the first point, we limited analysis to symptomatic patients because all asymptomatic patients were not reported as symptomatic patients. We also implicitly assumed that asymptomatic patients and symptomatic patients were the same in terms of infectiousness. Therefore, we must check that point.

Thirdly, our proposed and used estimation procedure for the effective reproduction number might be experimental. Its robustness should be examined as the next challenge for future research.

Fourthly, the source of infection and infected place were identified through contact tracing by the public health center. It was not based on the lineage of the genetic sequence of SARS-CoV-2 or other method. Therefore, we must remark that this information might include some ambiguity.

## Conclusion

We demonstrated that effective reproduction numbers from children were very low and that they were higher among adults and elderly people than among the same age class. Moreover, although the highest and second-highest places of most frequent infections were “other” and at “home” with some exceptions, infection at “hospitals” was remarkable among adult and elderly people. Among elderly people, infection at facilities for elderly people was also high. The relative frequencies of infections at nursing schools, schools, restaurants, and entertainment nightspots were negligible.

The present study is based on the authors’ opinions: it does not reflect any stance or policy of their professionally affiliated bodies.

## Data Availability

Japan Ministry of Health, Labour and Welfare. Press Releases.

https://www.mhlw.go.jp/stf/newpage_10723.html

## Acknowledgments

We acknowledge the great efforts of all staff at public health centers, medical institutions, and other facilities who are fighting the spread and destruction associated with COVID-19.

## Ethical considerations

All information used for this study was collected under the Law of Infection Control, Japan and published data was used. There is therefore no ethical issue related to this study.

## Competing Interest

No author has any conflict of interest, financial or otherwise, to declare in relation to this study.

## Notes

### Competing Interest Statement

The authors have declared no competing interest.

### Funding Statement

The author(s) received no specific funding for this work.

### Author Declarations

All information used for this study has been published elsewhere. There is therefore no ethical issue related to this study.

